# Relative reliability, construct validity, and concurrent validity of the Jiu-Jitsu Anaerobic Performance Test (JJAPT)

**DOI:** 10.64898/2026.06.24.26356347

**Authors:** Flávio Aurélio Fernandes Soares, Leandro Vidal Andreato, Hugo Enrico Souza Machado, Victor Silveira Coswig, Aldo Ângelo Moreira Lima

**Author notes:** **Corresponding author:** Flávio Aurélio Fernandes Soares, Postgraduate Program in Medical Sciences - Federal University of Ceará (UFC). Fortaleza, Ceará, Brazil.

## Abstract

This study aimed to investigate the test-retest reliability of the Jiu-Jitsu Anaerobic Performance Test (JJAPT), as well as its construct validity according to experience level (intermediate and advanced) and belt rank (blue, purple, brown, and black), in addition to its concurrent validity with physical performance measures. Twenty-three amateur Brazilian Jiu-Jitsu (BJJ) athletes participated in two assessment sessions. JJAPT repetitions (mean, peak, and total) were analyzed under 4 and 5-set protocols, along with physical tests including vertical jump, Wingate test, and specific strength-endurance tests. Reliability was assessed using the intraclass correlation coefficient (ICC), construct validity through t-tests and ANOVA, and concurrent validity through Pearson’s correlation. Results indicated that the JJAPT showed moderate to good reliability (ICC = 0.74 to 0.82). No significant differences were observed between experience levels or belt ranks for any variables (p > 0.05), with trivial effect sizes, indicating a lack of discriminant validity. On the other hand, significant correlations were found between JJAPT performance and isometric and dynamic strength-endurance tests (r = 0.39 to 0.62), with moderate to large magnitudes. It is concluded that the JJAPT shows measurement stability; however, it demonstrates limited ability to discriminate performance between athletes of different experience levels and belt ranks, being more strongly associated with strength-endurance capacities.

## INTRODUCTION

Brazilian jiu-jitsu (BJJ) is a combat sport whose primary objective is to submit opponents through chokeholds and joint-lock techniques (1). Furthermore, BJJ is characterized as an intermittent, high-intensity combat sport in which physiological and neuromuscular demands, such as energy resynthesis and force production, are substantial to support technical actions throughout the match, which may last up to 10 minutes (2). Therefore, optimizing the physical capacities of a BJJ athlete represents a fundamental component of the training process (3). These physiological and neuromuscular demands require substantial contributions from both the aerobic and anaerobic energy systems for energy supply (4). In this context, although the oxidative system predominates in energy resynthesis during matches across different combat sports, decisive technical actions are predominantly performed rapidly and at high-intensity and, consequently, rely primarily on anaerobic energy pathways (e.g., phosphagen and glycolytic systems) for energy provision (5–7).

Within this context, studies investigating the physiological responses in BJJ have been conducted using simulated match protocols (2,8), official competitions (9), and training sessions (10,11). During simulated no-gi (without kimono) BJJ matches, the glycolytic system demonstrates a substantial contribution to total anaerobic energy supply (i.e., 72.1 ± 11.9%), whereas the phosphagen system contributes approximately 27.9 ± 11.9% (12). It is important to note that analyses of the temporal structure of BJJ matches have demonstrated an effort:pause ratio of approximately 10:1, particularly characterized by actions and movements performed on the ground (13). Furthermore, when examining the temporal distribution of these actions, effort periods are composed of approximately 25-second low-intensity bouts (i.e., grip disputes and isometric force actions), interspersed with approximately 5 seconds of high-intensity actions (i.e., takedown attempts, submission attempts, and guard passes) (14). These temporal characteristics reinforce the high anaerobic demand required to sustain high-intensity actions and highlight the importance of assessing this capacity in BJJ athletes.

Alternatively to general assessments (i.e., cycling Wingate; running RAST), sport-specific tests, considering movement patterns and physiological demands, have been suggested to provide a more specialized understanding of parameters related to actual sport performance (15). Therefore, to be considered valid, a performance test must assess physical capabilities and skills related to combat performance, replicating physical and physiological responses like those observed during combat situations (16). In this context, Villar et al. (2018) proposed the Jiu-Jitsu Anaerobic Performance Test (JJAPT) to assess anaerobic performance of BJJ athletes. The test consists of quantifying the number of repetitions performed across four or five 1-minute rounds interspersed with 45 seconds of recovery, based on a specific Jiu-Jitsu movement pattern (i.e., the “butterfly guard”) (17). The authors suggested that the JJAPT demonstrates a high capacity to reproduce the physiological demands of simulated matches; however, the claim that such similarity in responses constitutes evidence of the test’s effectiveness may be questioned.

In this sense, although the JJAPT was designed to assess anaerobic performance (Villlar at al., 2018), the duration of each set (1 min) and the total duration of the test (8 min) represent a clear limitation for anaerobic assessment. Furthermore, the test involves only one fighting situation (sweeps), without encompassing other frequent actions (e.g., takedowns and guard passes), which may also be considered a limitation. Despite this, previous studies have demonstrated that the JJAPT was able to discriminate performance between intermediate (blue and purple belts) and advanced athletes (brown and black belts) (18,19), regardless of their technical profile (guard players or guard passers) (18) or its correlation with other neuromuscular, aerobic, or anaerobic assessments (19). However, the competitive structure of official tournaments is organized through matchups between athletes of the same belt rank (20), which limits the ecological validity of the previously adopted study designs. In this context, it remains unclear whether the JJAPT demonstrates validity to discriminate performance among athletes of the same belt rank.

Therefore, it becomes relevant to investigate the validity of the JJAPT in discriminate intermediate (blue and purple belts) and advanced (brown and black belts) athletes) by their performance. The proposal is to verify the sensitivity of the test under conditions that more ecologically reflect the competitive structure and demands of the sport. Accordingly, this study aimed to investigate the test–retest reliability, as well as the construct and concurrent validity of JJAPT repetitions in amateur BJJ athletes.

## MATERIALS AND METHODS

### Participants

Twenty-three amateur BJJ athletes (20 males and 3 females) with 8.7±6.3 years of training experience, a mean age of 29.3±6.8 years, body mass of 78.7±14.6 kg, and height of 172±0.07 cm voluntarily agreed to participate in the study. The term “amateur athlete” refers to individuals who regularly participate in training sessions and competitions but do not consider the sport as their primary professional occupation (21). Participants were grouped according to experience level: intermediate (blue and purple belts) and advanced (brown and black belts), and belt rank: blue belt (n = 5), purple belt (n = 6), brown belt (n = 4), and black belt (n = 8), according to the classification established by the International Brazilian Jiu-Jitsu Federation (IBJJF) (20). All athletes met the inclusion criteria: (i) being physically able to perform the physical tests, (ii) having won medals in official competitions within the previous 6 months, and (iii) having at least 1 year of training experience. Athletes who did not complete the physical tests were excluded. All participants were informed about the study procedures, potential risks and benefits associated with participation, and their right to withdraw from the study at any time. The study was approved by the Ethics Committee of the Federal University of Ceará (protocol number: 6082118 e CAAE 67616923.7.0000.5054).

### Study design

The athletes were evaluated on two separate occasions (test and retest). During the first session, anthropometric measurements (body mass and height), a training and competition history questionnaire, and the JJAPT were administered. During the second session, athletes performed:

(1) vertical jump assessments; (2) anaerobic lower-limb power assessment (Wingate Test); (3) isometric (KGSTiso) and dynamic (KGSTdin) strength-endurance tests; and (4) the sport-specific anaerobic test (JJAPT). In both sessions, athletes were previously instructed to refrain from physical exercise within the 24 hours preceding testing, abstain from alcohol consumption, and maintain their habitual sleep routine. Before the beginning of the assessments, all participants performed 5 minutes of static stretching followed by a warm-up consisting of 5 minutes of low-intensity walking at a self-selected pace. A 5-minute interval was provided between tests to optimize recovery and minimize the effects of residual fatigue. All assessments were conducted between 11:00 a.m. and 1:00 p.m. at the Biomechanics Laboratory of the Institute of Physical Education and Sports of the Federal University of Ceará (IEFES/UFC).

### Anthropometrics assessments and training background

Body mass and height were assessed using a digital scale with an attached stadiometer (Sanny^®^, Brazil), with a maximum capacity of 200 kg (precision of 100 g) and a measurement range from 115 to 210 cm (precision of 1 cm). Thereafter, a self-reported questionnaire developed by the researchers was administered to collect information regarding the frequency and duration of training sessions. Subsequently, weekly training volume (TV) was calculated as the product of session duration (in minutes) and the number of training sessions performed per week.

### Countermovement Jump (CMJ) and Squat Jump (SJ)

The CMJ and SJ tests were performed using a contact mat system (Jump System Pro, Cefise^®^). To estimate jump height, the contact mat uses a timing system that begins recording when the participant loses contact with the platform with both feet and ends when the first foot touches the surface again. Based on the recorded flight time, the device automatically calculates jump height in centimeters (22).

The testing protocol consisted of three consecutive jump attempts, with a 5-second interval between jumps. For the CMJ, athletes stood upright on the contact mat with their hands fixed on the hips, knees extended, and trunk upright. Upon the evaluator’s signal, athletes were instructed to rapidly perform a knee and hip flexion movement (eccentric phase), followed by a rapid extension of the knees and hips (concentric phase), aiming to jump as high as possible. For the SJ, athletes performed the same procedure; however, a 2-second pause was required during the transition between the eccentric and concentric phases. The mean (CMJmean and SJmean) and peak values (SJpeak and CMJpeak) obtained from the three attempts were used for analysis. Test reliability demonstrated classifications ranging from good reliability for CMJmean (ICC₃,₁ = 0.86), SJmean (ICC₃,₁ = 0.83), and SJpeak (ICC₃,₁ = 0.82), to excellent reliability for CMJpeak (ICC₃,₁ = 0.93) (23). Additionally, mean (CMJmean/SJmean) and peak (CMJpeak/SJpeak) ratios were calculated to assess the eccentric utilization ratio as an indicator of the stretch-shortening cycle efficiency of the muscle–tendon system (24).

### Wingate Test

The Wingate test was performed using a Biotec 2100 cycle ergometer (Cefise®). After 5 minutes of low-intensity pedaling followed by 3 minutes of rest, a load adjusted according to the athlete’s body mass (0.075 kg·kg⁻¹) was applied to the cycle ergometer (25). Athletes were instructed to pedal as fast as possible during a single uninterrupted 30-second trial. Verbal encouragement was provided throughout the test to motivate the athletes. Data were processed using Ergometric 6.0 software, and values of relative peak power (PP), mean power (MP), minimum power (Pmin), and fatigue index (FI) were calculated.

### Kimono Grip Strength Test *– KGST*

Isometric (KGSTiso) and dynamic (KGSTdyn) strength-endurance were assessed using the Kimono Grip Strength Test (KGST) (26,27). A gi was positioned over a horizontal bar at a height of approximately 2 meters. For the KGSTiso, athletes were required to support their body mass for as long as possible (in seconds) while gripping the gi with both hands and maintaining maximal elbow flexion. For the KGSTdyn, the same testing setup was applied, with athletes performing the maximum number of repetitions, starting from a fully flexed elbow position to a fully extended elbow position. A single trial was allowed for each athlete to perform the test until voluntary exhaustion. The results were expressed as relative values normalized to body mass.

### Jiu-Jitsu Anaerobic Performance Test (JJAPT)

The JJAPT consists of performing the maximum number of repetitions of the “butterfly guard” technical movement across four or five 1-minute sets, interspersed with 45 seconds of recovery (17). The starting position consisted of the athlete lying in the supine position, with the knees flexed at approximately 45° and the feet hooked between the partner’s legs (butterfly guard position). The partner was required to present a body mass similar to that of the athlete, remain kneeling with an upright trunk and arms apart, and adopt a passive posture throughout the execution of the test. The athlete initiated the movement by flexing the hips and trunk until reaching a seated position, at which point the athlete wrapped both arms around the partner’s trunk near the axillary region. Subsequently, the athlete moved backward while lifting the partner using the butterfly guard until the partner’s head surpassed the guard line. Finally, the entire movement was reversed, returning the partner to the mat and resuming the initial position. At the end of each set, athletes reported their rating of perceived exertion using an adapted Borg scale ranging from 0 to 10, with corresponding descriptors (28). The mean (JJAPTmean), peak (JJAPTpeak), and total number of repetitions (JJAPTsum) obtained during the 4 and 5-set protocols were considered for analysis, according to the following equation:

JJAPTmean = total number of repetitions performed across all sets / number of sets. JJAPTpeak = highest number of repetitions achieved among the sets.

JJAPTsum = total number of repetitions performed across all sets.

### Statiscal Analysis

Data are presented as mean ± standard deviation. Data normality was verified using the Shapiro–Wilk test, and homogeneity of variance was assessed using the Levene test. The Intraclass Correlation Coefficient (ICC) was used to evaluate the relative test–retest reliability of the mean (JJAPTmean), peak (JJAPTpeak), and total (JJAPTsum) number of repetitions. ICC values were interpreted as follows: < 0.50 (poor reliability); 0.50–0.75 (moderate reliability); 0.75–0.90 (good reliability); and > 0.90 (excellent reliability) (23).

An independent samples t-test was used to examine differences in repetitions obtained during the retest between intermediate athletes (blue and purple belts) and advanced athletes (brown and black belts) for mean (JJAPTmean), peak (JJAPTpeak), and total repetitions (JJAPTsum). Effect size was determined using Cohen’s d and interpreted as follows: < 0.20 (trivial); 0.20–0.49 (small); 0.50–0.79 (moderate); and ≥ 0.80 (large)^1^. A one-way ANOVA was performed to analyze differences in retest repetitions among belt ranks (blue, purple, brown, and black belts) for the same JJAPT parameters. Effect size was calculated using omega squared (ω²), adopting the following thresholds: 0.00–0.01 (trivial); 0.01–0.05 (small); 0.06–0.13 (moderate); and ≥ 0.14 (large) (29).

Pearson’s correlation test was used to examine concurrent validity between the physical tests and the mean (JJAPTmean), peak (JJAPTpeak), and total number of repetitions (JJAPTsum). Hopkins’ classification (30) was adopted to interpret effect size magnitude, considering the following thresholds: 0.0–0.1 (trivial); 0.1–0.3 (small); 0.3–0.5 (moderate); 0.5–0.7 (large); 0.7–0.9 (very large); and 0.9–1.0 (nearly perfect). Statistical analyses were performed using JASP, adopting a significance level of 5% (p < 0.05)

## RESULTS

### 1 Sample Characterization and Comparison of Age, Anthropometric Variables and Training Volume

Age, anthropometric variables, and training volume were analyzed among athletes with different experience levels (intermediate and advanced) and belt ranks (blue, purple, brown, and black belts). The results demonstrated statistically significant differences according to experience level. Advanced athletes presented higher values for all variables, including age (t(21) = 3.49; MD = 8 years; p = 0.00), body mass (t(21) = 2.52; MD = 14 kg; p = 0.02), height (t(21) = 3.10; MD = 0.08 m; p = 0.00), and training volume (t(21) = 2.30; MD = 86 minutes; p = 0.03).

Among athletes of different belt ranks, the results demonstrated statistically significant differences for age (F(3,19) = 4.10; p = 0.02), body mass (F(3,19) = 3.90; p = 0.02), and height (F(3,19) = 6.17; p = 0.00). Bonferroni post hoc analysis indicated differences between black belt and purple belt athletes for age (MD = 9.62 years; SE = 3.08; 95% CI: 0.55 to 18.69; p = 0.03), body mass (MD = 22.26 kg; SE = 6.70; 95% CI: 2.52 to 42.00; p = 0.02), and height (MD = 0.12 m; SE = 0.03; 95% CI: 0.03 to 0.21; p = 0.00). Training volume did not demonstrate statistically significant differences according to belt rank (F(3,19) = 2.44; p = 0.09).

### 2 JJAPT Test Reliability

Test reliability demonstrated classifications ranging from “moderate” to “good” for the mean (JJAPTmean), peak (JJAPTpeak), and total number of repetitions (JJAPTsum) in both the 4- and 5-set protocols. The analysis indicated that the test is capable of producing consistent results across repeated assessments performed under the same conditions at different time points (Table 1).

**TABLE 1.** Intraclass Correlation Coefficient (ICC) results for the JJAPT in the 4- and 5-set protocols.

| 4 SETS | TEST | RETEST | ICC | IC95% | CLASSIFICATION |
| --- | --- | --- | --- | --- | --- |
| JJAPT<sub>mean</sub> | 17.6 ± 2.4 | 18.6 ± 2.1 | 0.82 | 0.63 – 0.92 | *Good* |
| JJAPT<sub>peak</sub> | 18.7 ± 2.4 | 19.6 ± 2 | 0.74 | 0.48 – 0.88 | *Moderate* |
| JJAPT<sub>sum</sub> | 69.6 ± 9.3 | 74 ± 8.9 | 0.82 | 0.63 – 0.92 | *Good* |
| 5 SETS | TEST | RETEST | ICC | IC95% | CLASSIFICATION |
| JJAPT<sub>mean</sub> | 17.5 ± 2.3 | 18.6 ± 2.2 | 0.75 | 0.49 – 0.88 | *Good* |
| JJAPT<sub>peak</sub> | 19 ± 2.4 | 20 ± 2.2 | 0.79 | 0.46 – 0.90 | *Good* |
| JJAPT<sub>sum</sub> | 88.2 ± 11.8 | 92.8 ± 11.1 | 0.80 | 0.58 – 0.90 | *Good* |

**JJAPT<sub>mean</sub>:** Repetitions mean. **JJAPT<sub>peak</sub>:** Peak repetition. **JJAPT<sub>sum</sub>:** Sum of the repetitions. **IC95%:** Confidence interval - 95%.

### 3 Construct validity of the JJAPT

The analysis of the JJAPT’s ability to discriminate between intermediate and advanced athletes did not demonstrate statistically significant differences for mean (JJAPTmean), peak (JJAPTpeak), or total repetitions (JJAPTsum) in either the 4 or 5-set protocols. Additionally, effect sizes were trivial (d < 0.2), indicating a low magnitude of differences between athletes with different experience levels. Similarly, when considering athletes across different belt ranks, no statistically significant differences were observed for any of the evaluated parameters (mean, peak, and total repetitions) in either protocol.

Tables 2 and 3 presents, respectively, the results of the statistical analyses according to experience level and belt rank for mean (JJAPTmean), peak (JJAPTpeak), and total repetitions (JJAPTsum).

**TABLE 2.** Comparison between belt ranks for the JJAPT in the 4- and 5-set protocols (mean ± standard deviation).

| 4 SETS | ADVANCED | INTERMEDIATE | Statistic t | p value | Cohen's d |
| --- | --- | --- | --- | --- | --- |
| JJAPT<sub>mean</sub> | 18 ± 2.29 | 18 ± 2.24 | -0.06 | *0.94* | *Trivial* |
| JJAPT<sub>peak</sub> | 19 ± 1.97 | 19 ± 2.21 | -0.46 | *0.64* | *Trivial* |
| JJAPT<sub>sum</sub> | 73 ± 8.80 | 74 ± 9.43 | -0.09 | *0.92* | *Trivial* |
| 5 SETS | ADVANCED | INTERMEDIATE | Statistic t | p value | Cohen's d |
| JJAPT<sub>mean</sub> | 18 ± 2.22 | 18 ± 2.42 | 0.12 | *0.90* | *Trivial* |
| JJAPT<sub>peak</sub> | 20 ± 2.36 | 19 ± 2.21 | 0.26 | *0.79* | *Trivial* |
| JJAPT<sub>sum</sub> | 93 ± 11.26 | 92 ± 11.52 | 0.27 | *0.78* | *Trivial* |

**JJAPT<sub>mean</sub>:** Mean number of repetitions. **JJAPT<sub>peak</sub>:** Peak repetition value. **JJAPT<sub>sum</sub>:** Total number of repetitions. **Cohen's d:** Cohen's classification for effect size.

**TABLE 3.**
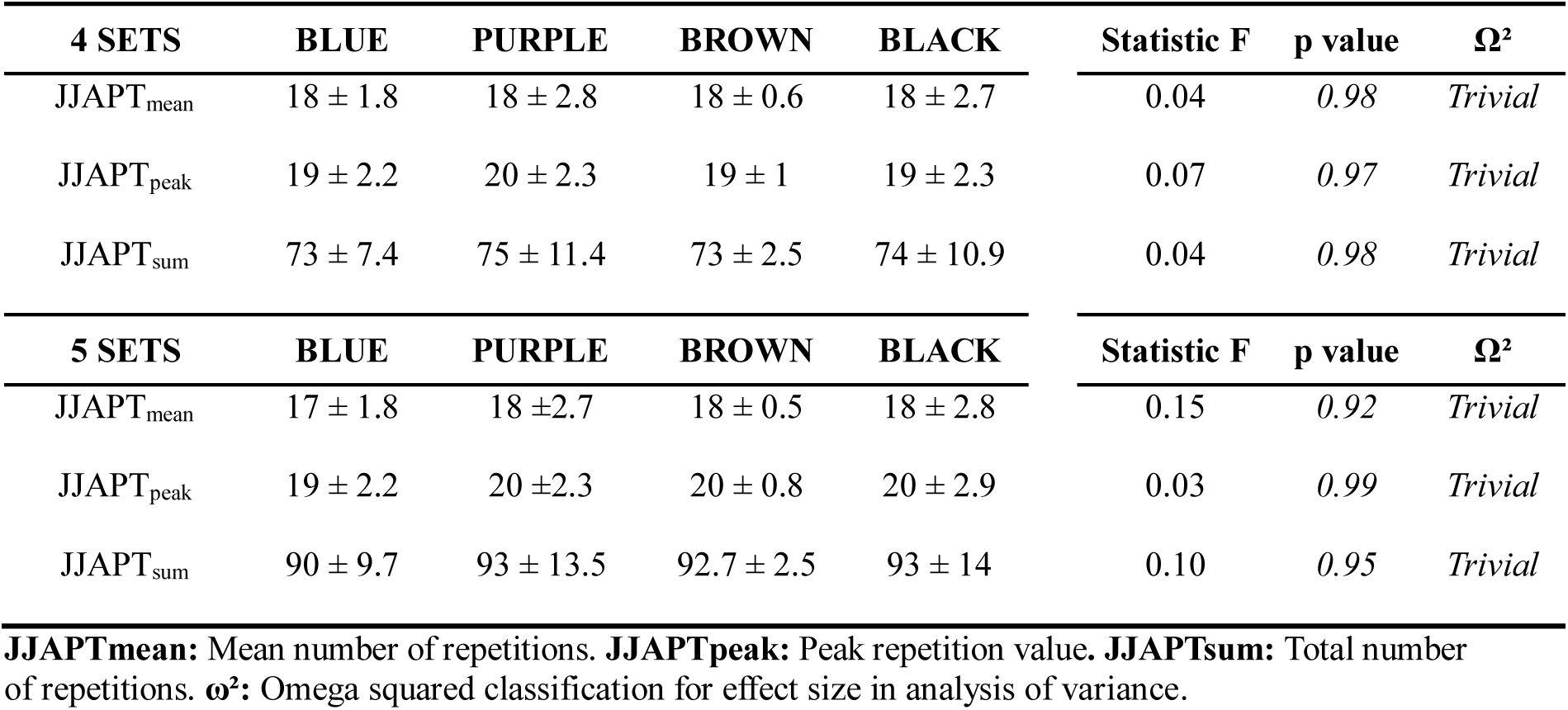
Comparison between belt ranks for the JJAPT in the 4- and 5-set protocols (mean ± standard deviation).

| 4 SETS | BLUE | PURPLE | BROWN | BLACK | Statistic F | p value | $\Omega^2$ |
| --- | --- | --- | --- | --- | --- | --- | --- |
| JJAPT<sub>mean</sub> | 18 ± 1.8 | 18 ± 2.8 | 18 ± 0.6 | 18 ± 2.7 | 0.04 | 0.98 | Trivial |
| JJAPT<sub>peak</sub> | 19 ± 2.2 | 20 ± 2.3 | 19 ± 1 | 19 ± 2.3 | 0.07 | 0.97 | Trivial |
| JJAPT<sub>sum</sub> | 73 ± 7.4 | 75 ± 11.4 | 73 ± 2.5 | 74 ± 10.9 | 0.04 | 0.98 | Trivial |
| 5 SETS | BLUE | PURPLE | BROWN | BLACK | Statistic F | p value | $\Omega^2$ |
| JJAPT<sub>mean</sub> | 17 ± 1.8 | 18 ± 2.7 | 18 ± 0.5 | 18 ± 2.8 | 0.15 | 0.92 | Trivial |
| JJAPT<sub>peak</sub> | 19 ± 2.2 | 20 ± 2.3 | 20 ± 0.8 | 20 ± 2.9 | 0.03 | 0.99 | Trivial |
| JJAPT<sub>sum</sub> | 90 ± 9.7 | 93 ± 13.5 | 92.7 ± 2.5 | 93 ± 14 | 0.10 | 0.95 | Trivial |

**JJAPT<sub>mean</sub>:** Mean number of repetitions. **JJAPT<sub>peak</sub>:** Peak repetition value. **JJAPT<sub>sum</sub>:** Total number of repetitions. **$\omega^2$ :** Omega squared classification for effect size in analysis of variance.

### 4 Concurrent validity between JJAPT and Physical Tests

The analysis of correlations between JJAPT variables and physical performance tests revealed positive correlations with both the isometric (KGSTiso) and dynamic (KGSTdin) strength-endurance tests. The magnitude of the correlations ranged from “moderate” to “large.” These results indicate that the number of repetitions performed during the JJAPT was directly proportional to both isometric strength-endurance time and the number of repetitions achieved in the dynamic strength-endurance test. Table 4 presents the correlation coefficients and effect sizes for the 4- and 5-set protocols.

**TABLE 4.** Correlation coefficients (r) between JJAPT and physical tests for concurrent validity.

| JJAPT 4 SETS PROTOCOL |  |  |  |  |  |  |
| --- | --- | --- | --- | --- | --- | --- |
|  | JJAPT<sub>mean</sub> | ES | JAPT<sub>pico</sub> | ES | JJAPT<sub>sum</sub> | ES |
| CMJ<sub>mean</sub> (cm) | 0.006<br>(0.97) | Trivial | 0.11<br>(0.6) | Small | 0.006<br>(0.97) | Trivial |
| CMJ<sub>peak</sub> (cm) | -0.10<br>(0.96) | Trivial | 0.09<br>(0.65) | Trivial | -0.01<br>(0.96) | Trivial |
| SJ<sub>mean</sub> (cm) | -0.03<br>(0.86) | Trivial | 0.06<br>(0.76) | Trivial | -0.03<br>(0.86) | Trivial |
| SJ<sub>peak</sub> (cm) | -0.003<br>(0.98) | Trivial | 0.09<br>(0.67) | Trivial | -0.003<br>(0.98) | Trivial |
| CMJ<sub>mean</sub>/SJ<sub>mean</sub> (cm) | 0.04<br>(0.84) | Trivial | 0.12<br>(0.57) | Small | 0.04<br>(0.84) | Trivial |
| CMJ<sub>peak</sub>/SJ<sub>peak</sub> (cm) | -0.04<br>(0.83) | Trivial | 0.02<br>(0.9) | Trivial | -0.04<br>(0.8) | Trivial |
| PP (W.Kg) | -0.01<br>(0.93) | Trivial | 0.11<br>(0.59) | Small | -0.01<br>(0.93) | Trivial |
| MP (W.Kg) | -0.16<br>(0.45) | *Trivial* | -0.01<br>(0.93) | *Trivial* | -0.16<br>(0.45) | *Trivial* |
| P<sub>min</sub> (W.Kg) | -0.05<br>(0.79) | *Trivial* | 0.01<br>(0.96) | *Trivial* | -0.05<br>(0.79) | *Trivial* |
| FI (%) | 0.02<br>(0.9) | *Trivial* | 0.05<br>(0.79) | *Trivial* | 0.02<br>(0.9) | *Trivial* |
| KGST<sub>dyn</sub> (Rept.Kg) | **0.56**<br>**(0.00)** | *Large* | **0.62**<br>**(0.00)** | *Large* | **0.56**<br>**(0.00)** | *Large* |
| KGST<sub>iso</sub> (Sec.Kg.) | **0.42**<br>**(0.04)** | *Moderate* | **0.54**<br>**(0.00)** | *Large* | **0.42**<br>**(0.04)** | *Moderate* |
| **JJAPT 5 SETS PROTOCOL** |  |  |  |  |  |  |
| CMJ<sub>mean</sub> (cm) | 0.04<br>(0.82) | *Trivial* | 0.12<br>(0.56) | *Small* | 0.009<br>(0.96) | *Trivial* |
| CMJ<sub>peak</sub> (cm) | 0.02<br>(0.9) | *Trivial* | 0.1<br>(0.63) | *Small* | -0.008<br>(0.97) | *Trivial* |
| SJ<sub>mean</sub> (cm) | 0.02<br>(0.93) | *Trivial* | 0.1<br>(0.64) | *Small* | -0.02<br>(0.9) | *Trivial* |
| SJ<sub>peak</sub> (cm) | 0.05<br>(0.81) | *Trivial* | 0.12<br>(0.55) | *Small* | 0.008<br>(0.97) | *Trivial* |
| CMJ<sub>mean</sub>/SJ<sub>mean</sub> (cm) | -0.004<br>(0.98) | *Trivial* | 0.04<br>(0.85) | *Trivial* | 0.01<br>(0.94) | *Trivial* |
| CMJ<sub>peak</sub>/SJ<sub>peak</sub> (cm) | -0.14<br>(0.51) | *Trivial* | -0.13<br>(0.54) | *Trivial* | -0.08<br>(0.68) | *Trivial* |
| PP (W.Kg) | 0.17<br>(0.43) | *Small* | 0.2<br>(0.35) | *Small* | 0.05<br>(0.81) | *Trivial* |
| MP (W.Kg) | 0.18<br>(0.39) | *Small* | 0.04<br>(0.83) | *Trivial* | -0.13<br>(0.54) | *Trivial* |
| P<sub>min</sub> (W.Kg) | 0.009<br>(0.96) | *Trivial* | 0.07<br>(0.72) | *Trivial* | 0.004<br>(0.98) | *Trivial* |
| FI (%) | 0.1<br>(0.63) | *Small* | 0.05<br>(0.82) | *Trivial* | 0.01<br>(0.94) | *Trivial* |
| KGST<sub>dyn</sub> (Rept.Kg) | **0.52**<br>**(0.01)** | *Large* | **0.61**<br>**(0.00)** | *Large* | **0.54**<br>**(0.01)** | *Large* |
| KGST<sub>iso</sub> (Sec.Kg.) | **0.39**<br>**(0.05)** | *Moderate* | **0.46**<br>**(0.02)** | *Moderate* | 0.39<br>(0.06) | *Moderate* |

**Bold:** Statistically significant ( $p < 0.05$ ). KGST - Kimono Grip Strength Test Isometric (KGST<sub>ISO</sub>) and Dynamic (KGST<sub>DYN</sub>). Wingate test Peak power (PP), mean power (MP), minimum power (P<sub>min</sub>), and fatigue index (FI). JJAPT<sub>mean</sub>: Mean number of repetitions. JJAPT<sub>peak</sub>: Peak repetition value. JJAPT<sub>sum</sub>: Total number of repetitions. W – watts. Sec – seconds. Rept – Repetitions.

## DISCUSSION

The aim of the present study was to investigate the reliability of the JJAPT, construct validity across experience levels (intermediate and advanced) and belt ranks (blue, purple, brown, and black), and concurrent validity with physical performance measures. The main findings indicated that the JJAPT i) demonstrated moderate to good reliability across two distinct assessment moments; ii) did not show discriminant validity between experience levels or belt ranks and; iii) exhibited significant correlations with isometric and dynamic strength-endurance tests. Although the JJAPT provides consistent and stable repetition-based measures, the absence of construct validity and its restricted association with strength-endurance capacities may limit its potential to discriminate athletes across different belt ranks.

The JJAPT demonstrated good reliability for most of the evaluated parameters (mean, peak, and total repetitions) in both protocols (4 and 5 sets). The exception was the peak repetition in the 4-set protocol, which showed moderate reliability. Nevertheless, these results indicate test stability across different time points, suggesting overall reliability of the measures. Reproducible tests reduce the influence of random error and allow potential changes in performance outcomes to be more confidently attributed to actual changes in athlete performance (16). In this context, although the previous study by da Silva et al. (2019) reported excellent reliability of the test, the analysis was limited to the total number of repetitions across the 5 assessment sets. In contrast, the present study expanded this perspective by demonstrating that not only the total sum, but also the mean and peak repetitions can be considered valid performance indicators. This suggests that the JJAPT may be used as a tool for monitoring sport-specific performance in BJJ athletes and may be capable of identifying responses to training interventions, fatigue, or recovery status. Furthermore, it is important to emphasize that, within sports sciences, assessment tools that offer practical and feasible testing procedures are strongly encouraged (31). The ease of application of the JJAPT, especially the four-set version as suggested in the original proposal, without the need for expensive or difficult-to-access equipment, provides a high cost-benefit ratio, enabling athletes and coaches to implement the test in practical training settings, making it a viable and cost-effective alternative.

However, in the present study, the test did not demonstrate strong construct validity. In other words, the JJAPT was not able to discriminate athletes according to experience level or belt rank, indicating that intermediate and advanced athletes may present similar performance outcomes, as well as black belt athletes may exhibit comparable performance to blue belt athletes, for example. In this context, the homogeneity of the sample may have reduced the statistical power of the test to detect differences in performance. Athletes competing at higher levels (e.g., international and Olympic level) typically present superior physical performance compared to their lower-level counterparts (e.g., regional and national level) (32,33). In this sense, despite differences in training volume between experienced and novice athletes, the relatively similar competitive level of the sample may have resulted in low performance variability. However, this interpretation should be made with caution, as the discriminative capacity of the JJAPT across athletes of different competitive levels has not yet been fully established. Furthermore, belt rank itself may not reflect linear differences in physical capacities, as athletes of different ranks may present similar anaerobic performance levels. This is because belt progression in BJJ is influenced by training experience and sport-specific cultural factors; therefore, differences in belt rank do not necessarily correspond to proportional differences in the physiological capacities assessed.

Another possible explanation is related to the ecological limitations of the test. Although the JJAPT is considered a sport-specific performance measure, it may not adequately reproduce the technical and tactical complexity of actual combat. This suggests that the mechanics of the movement, in isolation, may not be sufficient to ensure construct validity for the test. In studies involving other grappling modalities such as Judo, the application of sport-specific performance tests (i.e., the Special Judo Fitness Test – SJFT) has demonstrated good capacity to discriminate athletes of different performance levels (34–36). Likely, the technical component of the SJFT is considered more complex, as it involves displacement and the execution of a throwing technique. The more dynamic nature of the SJFT may reflect a broader range of physical capacities, in addition to technical aspects, when compared to the JJAPT (36). It is noteworthy that SJFT has a time-motion (effort:pause ratio) structure that is quite similar to Judo matches (2:1), which is not the case regarding JJAPT, and may influence the test validity.

Although the study by da Silva et al. (2019) demonstrated sensitivity when comparing novice (blue and purple belts) to advanced athletes (brown and black belts), potential differences in sample composition among the studies should be considered. Also, it is important to highlight that BJJ competitions are structured according to belt rank equivalence (20). In this sense, the present study reinforces that the JJAPT may not constitute a valid assessment tool for differentiating athletes based exclusively on belt rank. The absence of discrimination between ranks may indicate that performance in the JJAPT is more closely related to physical fitness responses than to the athletes’ technical level, a finding that is partially supported by the lack of differences observed between guard players and passers in the test outcomes (18).

The anaerobic system constitutes the primary energy pathway recruited during high-intensity and short-duration efforts (37), as required by the technical actions performed during a BJJ match (1). The improvement of glycolytic capacity for energy production results from metabolic and musculoskeletal adaptations (38–40) in response to these stimuli. Thus, the stability of the protocol may allow temporal physiological adaptations, such as strength endurance, to be observed through the test over a training period.

In the present study, the mean, peak, and total repetitions in the JJAPT showed significant correlations with upper-limb strength-endurance tests (dynamic and isometric) performed under sport-specific conditions using the gi. Also, our findings revealed that JJAPT was not associated with any of the Wingate test variables (considered a gold standard for assessing total anaerobic capacity), which is in accordance with a previous study (19). This result may be considered novel, as it suggests that the JJAPT may assess a different component than originally proposed. It is likely that the JJAPT is more closely associated with specific neuromuscular capacities, such as strength endurance. From a mechanical standpoint, both the JJAPT and the dynamic and isometric gi-specific strength-endurance tests share similarities, including repetitive actions, tolerance to peripheral fatigue, and high levels of localized muscular endurance (26).

Additionally, the absence of correlations between jumping variables and the JJAPT in the present study suggests that performance in the test does not appear to depend substantially on lower-limb power. Considering that jump tests predominantly assess muscular power and rapid force production (41), these findings reinforce the idea that the JJAPT may be more closely related to muscular endurance. Taken together, these results support the hypothesis that the JJAPT may not exclusively represent total anaerobic capacity, but rather a construct that is primarily associated with sport-specific strength endurance.

Finally, the present study has some limitations that should be considered when interpreting the results. The relatively small sample size and the homogeneity of participants’ competitive level may have limited the JJAPT’s discriminative capacity across different belt ranks and experience levels. Therefore, future studies should include larger samples and athletes with different competitive levels (e.g., regional, national, and international), in order to increase performance variability and further investigate the test’s discriminant validity. In addition, future research should consider longitudinal designs to verify the JJAPT’s sensitivity to training-induced adaptations. Comparing responses across different training phases may help determine the test’s ability to monitor meaningful physical and physiological changes relevant to BJJ performance, given that the JJAPT presents reproducible measures.

## CONCLUSION

The Jiu-Jitsu Anaerobic Performance Test (JJAPT) showed moderate to good reliability, which may support its potential use for performance monitoring. However, it did not demonstrate discriminant validity for differentiating athletes by experience level or belt rank. The JJAPT correlated significantly with isometric and dynamic strength-endurance tests, suggesting it may reflect muscular endurance and fatigue tolerance more than anaerobic capacity. Overall, its use for distinguishing competitive level should be interpreted with caution.

## Data Availability

All data produced in the present study are available from the corresponding author upon reasonable request.

## Acknowledgments

The authors thank the athletes who participated in the study for their availability and collaboration throughout all stages of the research. They also thank the institutions and laboratories involved for the structural and scientific support provided for the study. This manuscript is original, has not been published previously, and is not under consideration elsewhere. All authors have approved the submission. The corresponding author holds active affiliation with the Federal University of Ceará (Brazil) and meets eligibility criteria under the CAPES Read & Publish agreement. Upon acceptance, we will select Open Access publication under the CC BY license.

## REFERENCES

1. Andreato LV, Follmer B, Celidonio CL, Honorato ADS. Brazilian Jiu-Jitsu Combat among Different Categories: Time-Motion and Physiology. A Systematic Review. Strength and Conditioning Journal. 2016;38(6):44–54. doi:10.1519/SSC.0000000000000256

2. Andreato LV, Marta S, Moraes F De, Victor J, Conti D, Regina R, et al. PHYSIOLOGICAL RESPONSES AND RATE OF PERCEIVED EXERTION IN BRAZILIAN JIU-JITSU ATHLETES. Kinesiology. 2012;44(2):173–81.

3. Jones NB, Ledford E. Strength and Conditioning for Brazilian Jiu-jitsu. Strength & Conditioning Journal. 2012 Apr;34(2):60. doi:10.1519/SSC.0b013e3182405476

4. Artioli GG, Bertuzzi RC, Roschel H, Mendes SH, Jr AHL, Franchini E. Determining the Contribution of the Energy Systems During Exercise. Journal of Visualized Experiments. 2012;61(March):3–7. doi:10.3791/3413

5. Campos FAD, Bertuzzi R, Dourado AC, Santos VGF, Franchini E. Energy demands in taekwondo athletes during combat simulation. Eur J Appl Physiol. 2012 Apr;112(4):1221–8. doi:10.1007/s00421-011-2071-4 PubMed PMID: 21769736.

6. Franchini E. Energy System Contributions during Olympic Combat Sports: A Narrative Review. Metabolites. 2023 Feb 17;13(2):297. doi:10.3390/metabo13020297 PubMed PMID: 36837916; PubMed Central PMCID: PMC9961508.

7. Julio UF, Panissa VLG, Esteves JV, Cury RL, Agostinho MF, Franchini E. Energy-System Contributions to Simulated Judo Matches [Internet]. 2017 May 1. doi:10.1123/ijspp.2015-0750

8. Andreato L V., Franchini E, De Moraes SMF, Pastório JJ, da Silva DF, Esteves JVDC, et al. Physiological and technical-tactical analysis in brazilian jiu-jitsu competition. Asian Journal of Sports Medicine. 2013;4(2):137–43. doi:10.5812/asjsm.34496 PubMed PMID: 23802056.

9. Diaz-Lara FJ, Del Coso J, García JM, Abián-Vicén J. Analysis of physiological determinants during an international Brazilian jiu-jitsu competition. International Journal of Performance Analysis in Sport. 2015;15(2):489–500. doi:10.1080/24748668.2015.11868808

10. Ribeiro R, Oliveira Silva JÍ, Dantas MGB, Menezes ES, Pereira Arruda AC, Schwingel PA. High-intensity interval training applied in Brazilian Jiu-jitsu is more effective to improve athletic performance and body composition. Journal of Combat Sports and Martial Arts. 2015;6(1):1–5. doi:10.5604/20815735.1166073

11. Rodrigues-Krause J, Silveira FP da, Farinha JB, Junior JV, Marini C, Fragoso EB, et al. Cardiorespiratory Responses and Energy Contribution in Brazilian Jiu-Jitsu Exercise Sets. International Journal of Performance Analysis in Sport. 2020;20(6):1092–106. doi:10.1080/24748668.2020.1829429

12. Pessôa Filho DM, Sancassani A, da Cruz Siqueira LO, Massini DA, Almeida Santos LG, Neiva CM, et al. Energetics contribution during no-gi Brazilian jiu jitsu sparring and its association with regional body composition. PLoS One. 2021;16(11):e0259027. doi:10.1371/journal.pone.0259027 PubMed PMID: 34767563; PubMed Central PMCID: PMC8589206.

13. Andreato LV, Julio UF, Gonçalves Panissa VL, Del Conti Esteves JV, Hardt F, Franzói de Moraes SM, et al. Brazilian Jiu-Jitsu Simulated Competition Part II: Physical Performance, Time-Motion, Technical-Tactical Analyses, and Perceptual Responses. J Strength Cond Res. 2015 Jul;29(7):2015–25. doi:10.1519/JSC.0000000000000819 PubMed PMID: 25559902.

14. Del Vecchio F, Stefania B, Hirata S, Chacon-Mikahi M. Análise morfo-funcional de praticantes de Brazilian jiu-jitsu e estudo da temporalidade e da quantificação das ações motoras na modalidade. MOVIMENTO e PERCEPÇÃO. 2007 Jan 1;7.

15. Vigh-larsen JF, Junge N, Cialdella-kam L, Tomás R, Young L, Krustrup P, et al. Testing in Intermittent Sports — Importance for Training and Performance Optimization in Adult Athletes. Med Sci Sports Exerc. 2024;56(8):1505–37. doi:10.1249/MSS.0000000000003442

16. Currell K, Jeukendrup AE. Validity, reliability and sensitivity of measures of sporting performance. Sports Med. 2008;38(4):297–316. doi:10.2165/00007256-200838040-00003 PubMed PMID: 18348590.

17. Villar R, Gillis J, Santana G, Pinheiro DS, Almeida ALRA. Association between anaerobic metabolic demands during simulated brazilian jiu-jitsu combat and specific jiu-jitsu anaerobic performance test. Journal of Strength and Conditioning Research. 2018;32(2):432–40. doi:10.1519/jsc.0000000000001536 PubMed PMID: 27379962.

18. da Silva JN, Kons RL, de Lucas RD, Detanico D. Jiu-Jitsu-Specific Performance Test: Reliability Analysis and Construct Validity in Competitive Athletes. Journal of Strength and Conditioning Research. 2019;36(1):174–9. doi:10.1519/JSC.0000000000003429 PubMed PMID: 31800472.

19. Silva Junior JN, Penteado dos Santos R, Kons RL, Gillis J, Caputo F, Detanico D. Relationship between a Brazilian Jiu-Jitsu specific test performance and physical capacities in experience athletes. Science and Sports. 2022;37(3):209.e1-209.e9. doi:10.1016/j.scispo.2021.03.009

20. IBJJF. Regulamento de Compeitões. 2024.

21. Araújo CGS, Scharhang J. Athlete: a working definition for medical and health sciences research. Scandinavian Journal of Medicine & Science in Sports. 2016;4–7. doi:10.1111/sms.12632

22. Pérez-Castilla A, García-Ramos A. Evaluation of the Most Reliable Procedure of Determining Jump Height During the Loaded Countermovement Jump Exercise: Take-Off Velocity vs. Flight Time. J Strength Cond Res. 2018 Jul 1;32(7):2025–30. doi:10.1519/jsc.0000000000002583 PubMed PMID: 29570575.

23. Koo TK, Li MY. A Guideline of Selecting and Reporting Intraclass Correlation Coefficients for Reliability Research. Journal of Chiropractic Medicine. 2016;15(2):155–63. doi:10.1016/j.jcm.2016.02.012 PubMed PMID: 27330520.

24. Kipp K, Krzyszkowski J, Heeneman J. Hip moment and knee power eccentric utilisation ratios determine lower-extremity stretch-shortening cycle performance. Sports Biomechanics. 2019;00(00):1–11. doi:10.1080/14763141.2019.1579854

25. Bar-Or O. The Wingate Anaerobic Test An Update on Methodology, Reliability and Validity. Sports Medicine: An International Journal of Applied Medicine and Science in Sport and Exercise. 1987;4(6):381–94. doi:10.2165/00007256-198704060-00001 PubMed PMID: 3324256.

26. Branco BHM, Diniz E, da Silva Santos JF, Shiroma SA, Franchini E. Normative tables for the dynamic and isometric judogi chin-up tests for judo athletes. Sport Sci Health. 2017 Apr 1;13(1):47–53. doi:10.1007/s11332-016-0331-8

27. Franchini E, Eduardo C, Souza B De, Urasaki R, Silva R, Oliveira F De, et al. Teste de resistência de força isométrica e dinâmica na barra com o Judogi. In: Anais do III Congresso de la Asociación Española de Ciencias del Deporte. 2004.

28. Nakamura FY, Moreira A, Aoki MS. Monitoramento da carga de treinamento: a percepção subjetiva do esforço da sessão é um método confiável? Revista da Educação Física/UEM. 2010;21(1):1–11. doi:10.4025/reveducfis.v21i1.6713

29. Olejnik S, Algina J. Generalized Eta and Omega Squared Statistics: Measures of Effect Size for Some Common Research Designs. Psychological Methods. 2003;8(4):434–47. doi:10.1037/1082-989X.8.4.434

30. Hopkins WG. A New View of Statistics. A scale of magnitudes for effect statistics. Sport Sci. 2002.

31. Machado HES, Santos Sousa JL, da Silva BS, Sant’Ana J, Diefenthaeler F, Coswig VS, et al. Reliability of aerobic fitness in combat sports: Insights from the ITStriker app specific incremental test. Proceedings of the Institution of Mechanical Engineers, Part P: Journal of Sports Engineering and Technology. 2025 May 24;1–8. doi:10.1177/17543371251343132

32. Campos BT, Abad CCC, Macedo Penna E, Lima COV, Avelar AF, Mara de Rezende C, et al. Reference Values for Maximal Strength in Elite Judo Athletes: National- and International-Level Competitors. Research Quarterly for Exercise and Sport. 2026 Apr 17;0(0):1–10. doi:10.1080/02701367.2026.2636255 PubMed PMID: 41996598.

33. Emerson F, Takito MY, Kiss MAPD, Sterkowicz S. Physical fitness and anthropometrical differences between elite and non-elite judo players. Biology of Sport. 2005;22(4):315–28.

34. Agostinho MF, Junior JAO, Stankovic N, Escobar-Molina R, Franchini E. Comparison of special judo fitness test and dynamic and isometric judo chin-up tests’ performance and classificatory tables’ development for cadet and junior athletes. J Exerc Rehabil. 2018 Apr 26;14(2):244–52. doi:10.12965/jer.1836020.010 PubMed PMID: 29740559; PubMed Central PMCID: PMC5931161.

35. Campos BT, Rodrigues JG da S, Machado HES, Coswig VS, Penna EM. DYNAMIC DISCRIMINATION: SJFT RESULT IN THE EVOLUTION OF STRENGTH AND POWER PERFORMANCE OF ELITE JUDO ATHLETES. J Phys Educ. 2025;36:e3601. 10.4025/jphyseduc.v36i1.3601

36. Sterkowicz-Przybycień K, Fukuda DH, Franchini E. Meta-Analysis to Determine Normative Values for the Special Judo Fitness Test in Male Athletes: 20+ Years of Sport-Specific Data and the Lasting Legacy of Stanisław Sterkowicz. Sports. 2019 Aug;7(8):194. doi:10.3390/sports7080194

37. Chamari K, Padulo J. ‘Aerobic’ and ‘Anaerobic’ terms used in exercise physiology: a critical terminology reflection. Sports Medicine - Open. 2015;1:9:1–4. doi:10.1186/s40798-015-0012-1

38. Abe T, Kitaoka Y, Kikuchi DM, Takeda K, Numata O, Takemasa T. High-intensity interval training-induced metabolic adaptation coupled with an increase in Hif-1 and glycolytic protein expression. J Appl Physiol. 2015;119:1297–302. doi:10.1152/japplphysiol.00499.2015

39. Mcginley XC, Bishop DJ. Influence of training intensity on adaptations in acid / base transport proteins, muscle buffer capacity, and repeated-sprint ability in active men. J Appl Physiol. 2016;121:1290–305. doi:10.1152/japplphysiol.00630.2016

40. Thomas ACQ, Stead CA, Burniston JG, Phillips SM. Exercise-specific adaptations in human skeletal muscle: Molecular mechanisms of making muscles fit and mighty. Free Radical Biology and Medicine. 2024;223(July):341–56. doi:10.1016/j.freeradbiomed.2024.08.010

41. Claudino JG, Cronin J, Mezêncio B, McMaster DT, McGuigan M, Tricoli V, et al. The countermovement jump to monitor neuromuscular status: A meta-analysis. Journal of Science and Medicine in Sport. 2017 Apr 1;20(4):397–402. doi:10.1016/j.jsams.2016.08.011

42. Cohen J. (1998). Statistical Power Analysis for the Behavioral Sciences (2nd ed.). Hillsdale, NJ: Lawrence Eribaum Associates, Publishers

